# Improving maternal postnatal check uptake in general practice using an opt-out equitable model of access: results of a 12-month quality improvement project

**DOI:** 10.1101/2025.09.06.25335222

**Authors:** Dhiviya Tharan

## Abstract

**BACKGROUND:** It is assumed that there is universal provision of the maternal postnatal 6-8 week check (6WC) in primary care following the introduction of additional funding provided through the General Medical Services contract in 2020/21. Prior to the pandemic, it is estimated that 20-40% of women in England did not have a postpartum maternal check recorded in primary care. Concerned that changes in local appointment access were contributing to an inequitable provision of postnatal care, we explored a model of access that improved the delivery of maternal postnatal care in general practice

**AIM:** To design a primary care model of access to improve the uptake of the maternal postnatal check that prioritised equitable access to care.

**DESIGN AND SETTING:** Cohort study and quality improvement project; women who had delivered a baby or stillborn delivery over 24 weeks gestation

**METHOD:** A retrospective pre-intervention clinical audit between April 2022 and March 2023 evaluated the service delivery performance of maternal postnatal 6WC. Implementation of a model of access with protected postnatal appointments and proactive invitation via SMS was introduced in April 2024. Post-intervention audit evaluated the intervention’s performance after 12 months.

**RESULTS:** Pre-intervention audit showed 58% (70/121) of eligible women had a maternal 6WC and 60% (42/70) were performed within 6–8 weeks after delivery. Following the introduction of the intervention, 98% (112/114) of eligible women were offered a postnatal check appointment. After 12 months, the uptake of maternal postnatal checks improved from 58% to 89% (101/114) and appointments performed within 6-8 weeks improving from 60% to 76% (77/101). The uptake of newborn checks improving from 86% to 91% (106/116) and appointments performed within 6-8 weeks improving from 46% to 75% (80/106).

**CONCLUSION:** We implemented protected postnatal appointments with proactive invitation via SMS and demonstrated a sustainable improvement in practice service delivery over 12 months of implementation. The protocol required no additional workforce resources, had a low administrative burden and used digital communication tools easily available to general practices nationwide. Our intervention provides a model of access for the provision of postnatal care in general practice to reduce inequality and inequity in healthcare.

## INTRODUCTION

Prior to the pandemic, it is estimated that 20-40% of women in England did not have a maternal postnatal 6-8 week check (6WC) recorded in primary care(1,2) and this reflects a global pattern(3,4). There are known disparities in the uptake of postnatal checks that negatively affects younger women and those in more deprived areas(2). There is an assumption that there is universal provision of the 6WC in primary care following the introduction of additional funding provided through the General Medical Services contract in 2020/21(5). Nationwide worsening maternal mortality statistics since the pandemic alongside significant ethnic disparities in outcomes as evidenced by the MBRRACE-UK study(6) suggests a lack of attention and investment in maternal health. National policies seek to redress disparities in women’s healthcare(7,8) by exploring healthcare access models which support equitable targeted approaches to improve outcomes by supporting improvements in access of care(9).

The COVID-19 pandemic disrupted the delivery of routine healthcare in general practice(10). A ‘supply-focussed’ model of access(11,12) was prioritised in policy interventions without comprehensive equality impact assessments and there is evidence that changes to delivery of healthcare compounded pre-existing health inequalities(13). Regional surveys called attention to the risks that general practice was not meeting women’s postnatal physical and mental health needs(14–16).

Concerned that changes in local appointment access were contributing to an inequitable provision of postnatal care, we proposed to evaluate our current model of access and rapidly innovate to implement an equity-inclusive model of access that improved the delivery of maternal postnatal care in our general practice.

## METHODOLOGY

Healthcare in the United Kingdom is provided through the publicly funded, comprehensive and universal National Health Service (NHS). Primary care is delivered by an independent GP contractor model. The responsibility of community postpartum care is shared between midwives, health visitors and general practitioners.

The project was undertaken in a general practice that serves the Stretford area in Trafford, a metropolitan borough of Greater Manchester, England. Trafford’s Index of Multiple Deprivation (IMD) Score in 2021 was 16.1 compared to the national average of 21.7(17). The practice population is approximately 14,000 patients.

The project aim was to meet the standard of care defined by the National Institute for Health and Care Excellence (NICE) Guidelines on Postnatal Care [NG194] that all eligible women undergo a postnatal check within 6-8 weeks after delivery(18).

The objectives of this project were to:

1. Measure the baseline uptake of the maternal six-week check (6WC) in one GP practice;
2. To design and deliver a model of access to improve the uptake of the maternal 6WC without compromising newborn 6WC provision and uptake;
3. And to measure the impact of the model on increasing uptake of the maternal 6WC.

The Six Sigma improvement process was used. A summary of the project is demonstrated in Figure 1.

**FIGURE 1.**
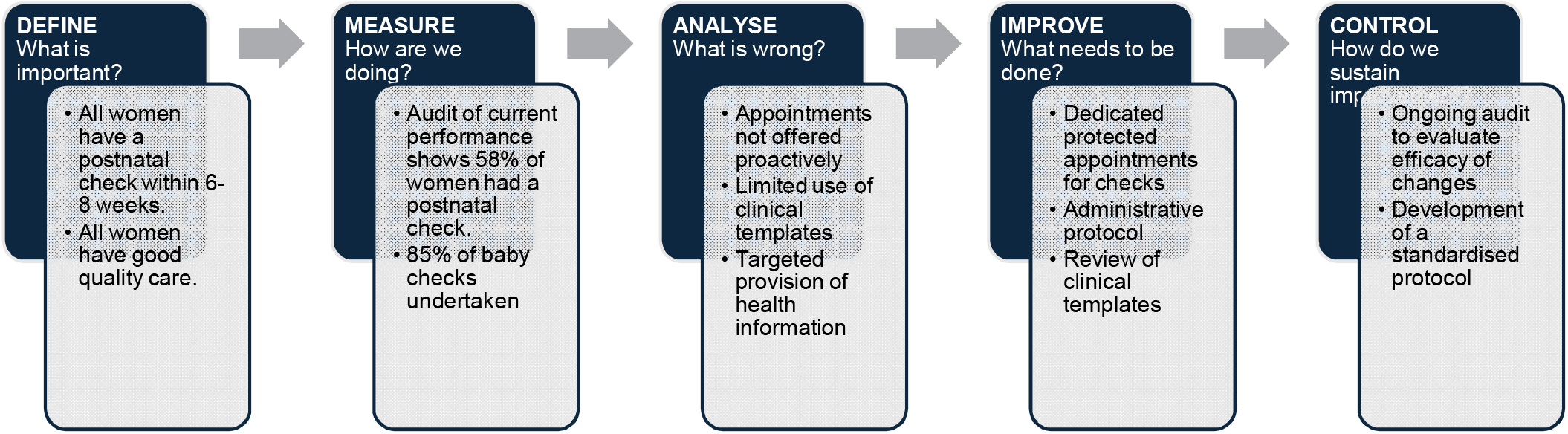

**FIGURE 2.**
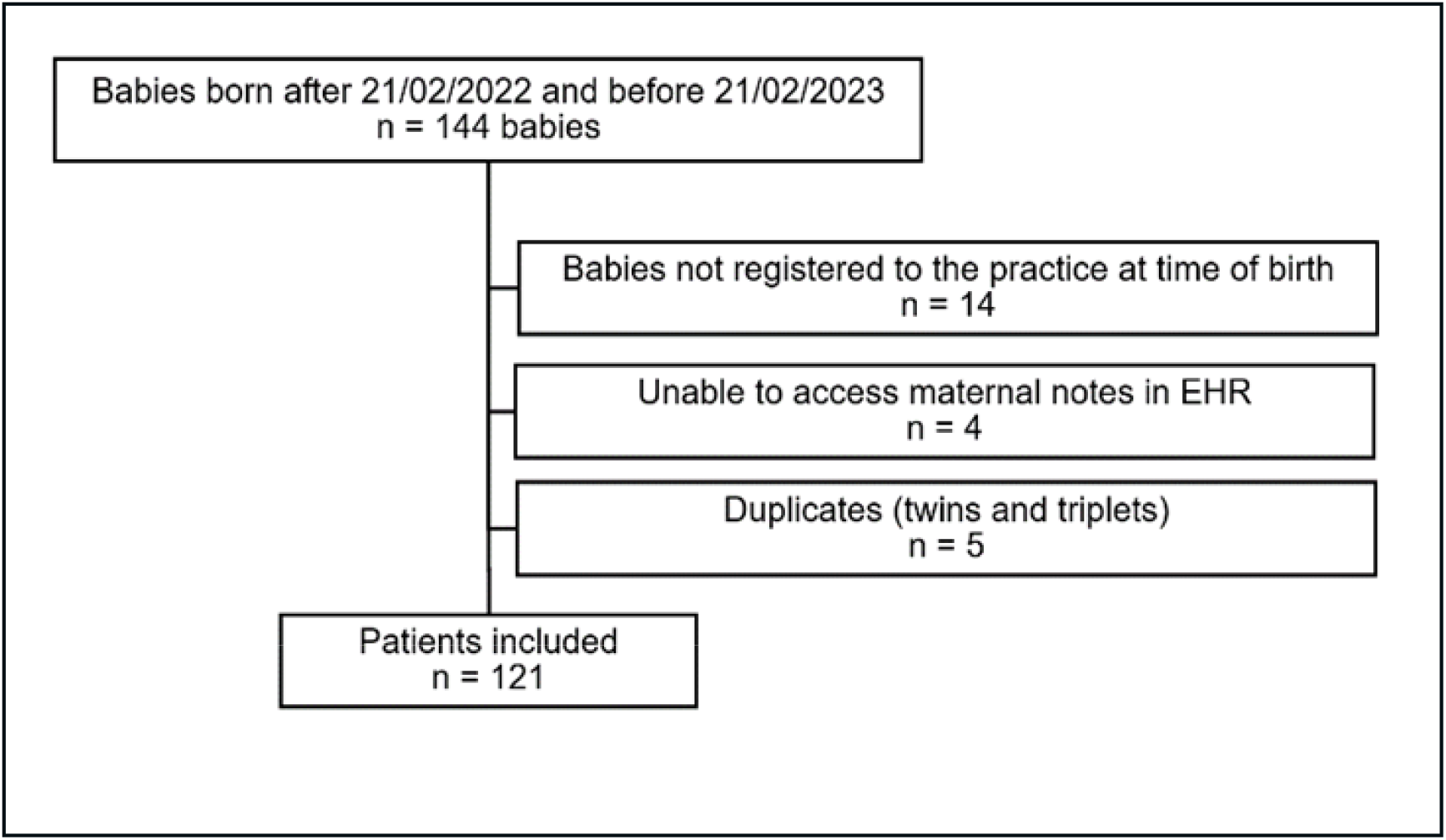

A retrospective clinical audit between April 2022 and April 2023 was undertaken to evaluate practice performance. During the audit period, the practice operated a digital-first patient-initiated model of access for unplanned clinical activity. The practice did not have a formal process for the delivery of postnatal checks to eligible women. Results were benchmarked against NICE Guidance on Postnatal Care [NG194] which recommend that every eligible woman should have a postnatal check 6-8 weeks after delivery(18).

Initial practice population searches reliant on clinical codes ([postnatal examination], [full postnatal examination], [postnatal maternal examination], [postnatal visits], [postpartum care], [Complete postnatal care], [[RFC] post-natal]) within our electronic health record (EHR) software, EMIS(19), produced an incomplete data set when compared to the estimated number of babies born within the same time period of February 2022 to February 2023.

To identify all eligible women, a patient search was run for “Babies born after 21/02/2022 and before 21/02/2023” on all registered patients in the practice. This identified 144 babies compared to the 29 women identified on searches dependent on clinical codes. Babies not registered to the practice at the time of their birth and babies where the maternal clinical notes could not be accessed were excluded. The baby’s mother was identified either via the “Household” information on EMIS or through Birth Certificate information. The mother’s patient record was reviewed for attendance at a postnatal appointment. Attendance at a postnatal appointment was recorded if a clinician used a postnatal check clinical code or, on reviewing clinical notes with no code, a systematic postnatal review of the patient was undertaken. Appointments that addressed a single issue (e.g. haemorrhoids, contraception prescriptions, mental health reviews) were not considered to be a comprehensive postnatal review. For individuals that did not have a postnatal check, appointments data was reviewed to determine if an appointment was offered. Checks completed within 40 – 58 days were accepted as being completed within ‘6-8 weeks’ to allow for flexibility in appointments arrangement. Ethnicity data, as self-reported by patients, were taken from patient records. Ethnicity data was not aggregated as recommended by Arrington et al.(20)

Data was collected by the author and input into and analysed using Microsoft Excel. A total of 121 patients eligible for postnatal checks were identified once multiple births were accounted for.

Between April 2022 and March 2023, 58% (70/121) of eligible women had a postnatal check and 60% (42/70) were performed within 6 – 8 weeks after delivery (see Tables 1 and 3). Ethnicity data was available for 53% (64/121) of the patient population (Table 2). Figure 3 displays the age distribution of audited patients. Evaluating babies registered to the practice at the time of birth, 86% (113/132) had a baby check and 46% (52/113) were performed within 6 – 8 weeks after birth.

**Table 1.** Population Data.

|  | *Pre-intervention population* |  |  |  | *Post-intervention population* |  |  |  |
| --- | --- | --- | --- | --- | --- | --- | --- | --- |
|  | Postnatal check performed |  | Postnatal check not performed |  | Postnatal check performed |  | Postnatal check not performed |  |
|  | n | % | n | % | n | % | n | % |
|  | 70 | 58% | 51 | 42% | 101 | 89% | 13 | 11 |
| **Age (years)** |  |  |  |  |  |  |  |  |
| Mean | 34 | - | 33 | - | 32 |  | 31 |  |
| Median | 34 | - | 32 | - | 33 |  | 32 |  |
| Age (range) | 23,46 | - | 18,44 | - | 18, 44 |  | 20, 38 |  |
| **Age (years)** |  |  |  |  |  |  |  |  |
| 18-22 | 0 |  | 3 |  | 4 |  | 2 |  |
| 23-27 | 7 |  | 8 |  | 19 |  | 1 |  |
| 28-32 | 28 |  | 18 |  | 35 |  | 4 |  |
| 33-37 | 27 |  | 11 |  | 32 |  | 5 |  |
| 38-42 | 6 |  | 10 |  | 10 |  | 1 |  |
| 43-47 | 2 |  | 1 |  | 1 |  | - |  |

**Table 2.** Population Ethnicity.

|  | *Pre-intervention population* |  | *Post-intervention population* |  |
| --- | --- | --- | --- | --- |
|  | Postnatal check performed (n) | Postnatal check not performed (n) | Postnatal check performed (n) | Postnatal check not performed (n) |
| **Ethnicity (self-reported)** |  |  |  |  |
| African |  |  | 2 |  |
| Asian or Asian British: other | 2 |  | 1 | 1 |
| Pakistani or British Pakistani | 3 | 3 | 9 | 3 |
| Bangladeshi or British Bangladeshi |  |  |  | 2 |
| Black: African | 2 | 1 | 1 |  |
| British or mixed British | 9 | 6 | 10 | 1 |
| Caribbean |  | 1 |  |  |
| Chinese | 1 |  | 1 |  |
| Indian or British Indian | 4 |  | 3 |  |
| Other |  |  | 2 |  |
| Other black background |  |  | 1 |  |
| Other mixed background | 2 |  |  |  |
| Other: Iranian |  | 1 |  |  |
| Polish |  | 1 |  |  |
| White and Black African |  |  | 1 |  |
| White and Black Caribbean |  |  | 1 |  |
| White British | 13 | 10 | 25 | 1 |
| White Irish |  | 1 |  |  |
| White Other | 3 |  | 2 | 1 |
| Ethnicity data not available | 30 | 27 | 42 | 5 |

**Table 3.** Intervention Outcomes.

|  | *Pre-intervention population* |  | *Post-intervention population* |  |
| --- | --- | --- | --- | --- |
|  | n | % | n | % |
| Checks completed within 42-56 days (strict) | 39 | 56% | 60 | 59% |
| Checks completed within 40-58 days (flexible) | 42 | 60% | 77 | 76% |
| Range: days to check | 32, 106 |  | 28, 94 |  |
| Average (mean) days to check | 55 |  | 51 |  |
| Average (median) days to check | 51 |  | 48 |  |

**Figure 3.**
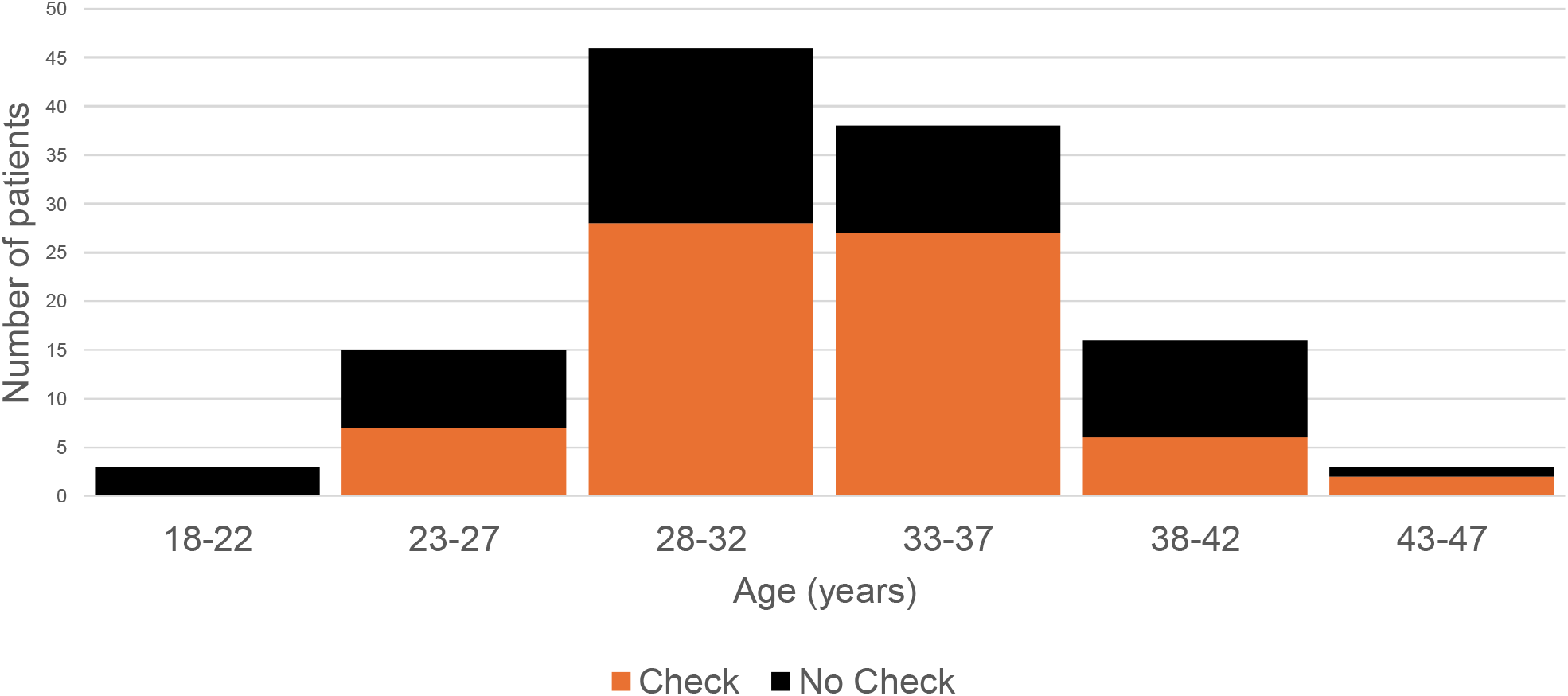
Histogram: Age distribution of pre-intervention population

The audit showed a suboptimal uptake of postnatal care and in those who did have a check, 60% were completed within the NICE recommendations of 6-8 weeks. Of the patients who did not have a postnatal check, 2/51 (4%) were pre-emptively offered an appointment indicating a need for an improved method of access to appointments.

Monthly data was analysed to understand practice demand over 12 months. Monthly appointment requirements ranged between 5 to 16 appointments per month (see Figure 4).

**Figure 4.**
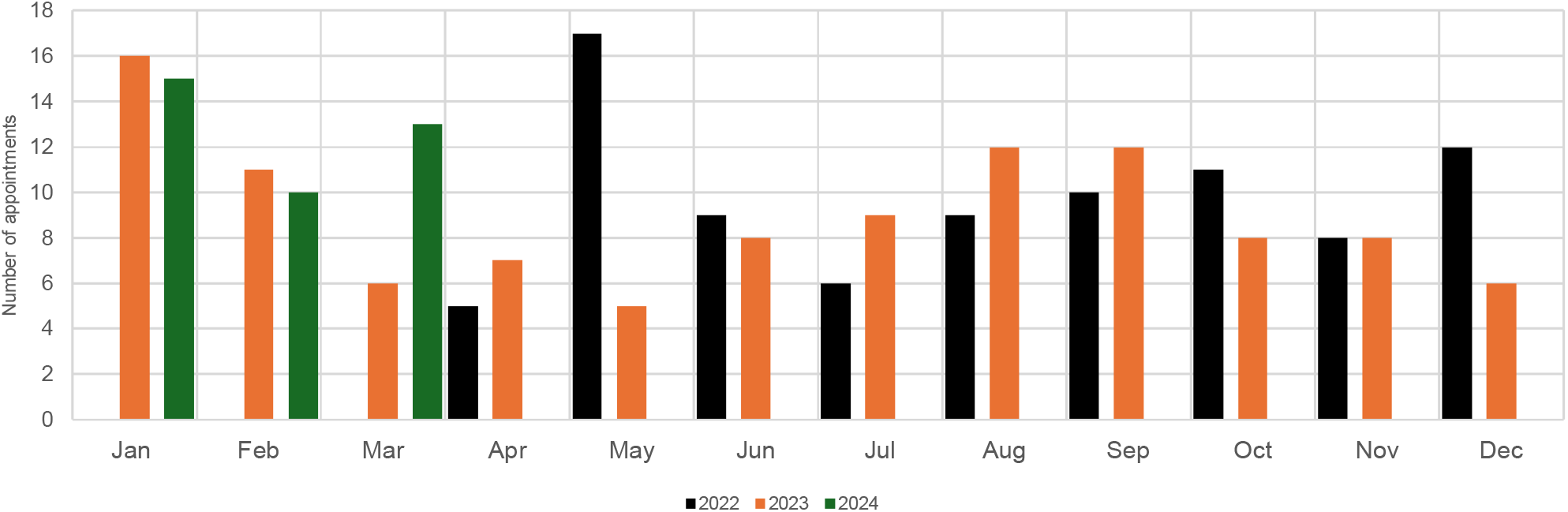
Monthly Practice Demand (2022-2024)

## INTERVENTION

The Greater Manchester Quality Improvement Framework identified six dimensions to be considered when approaching healthcare improvement(21). Following analysis of appointment data, we developed a patient-centred model of access that prioritised equity to all eligible patients that could be promptly deployed. As rapid innovation, efficiency and sustainability was required, we explored adaptations to our established clinical processes, as outlined in Figure 5.

**FIGURE 5.**
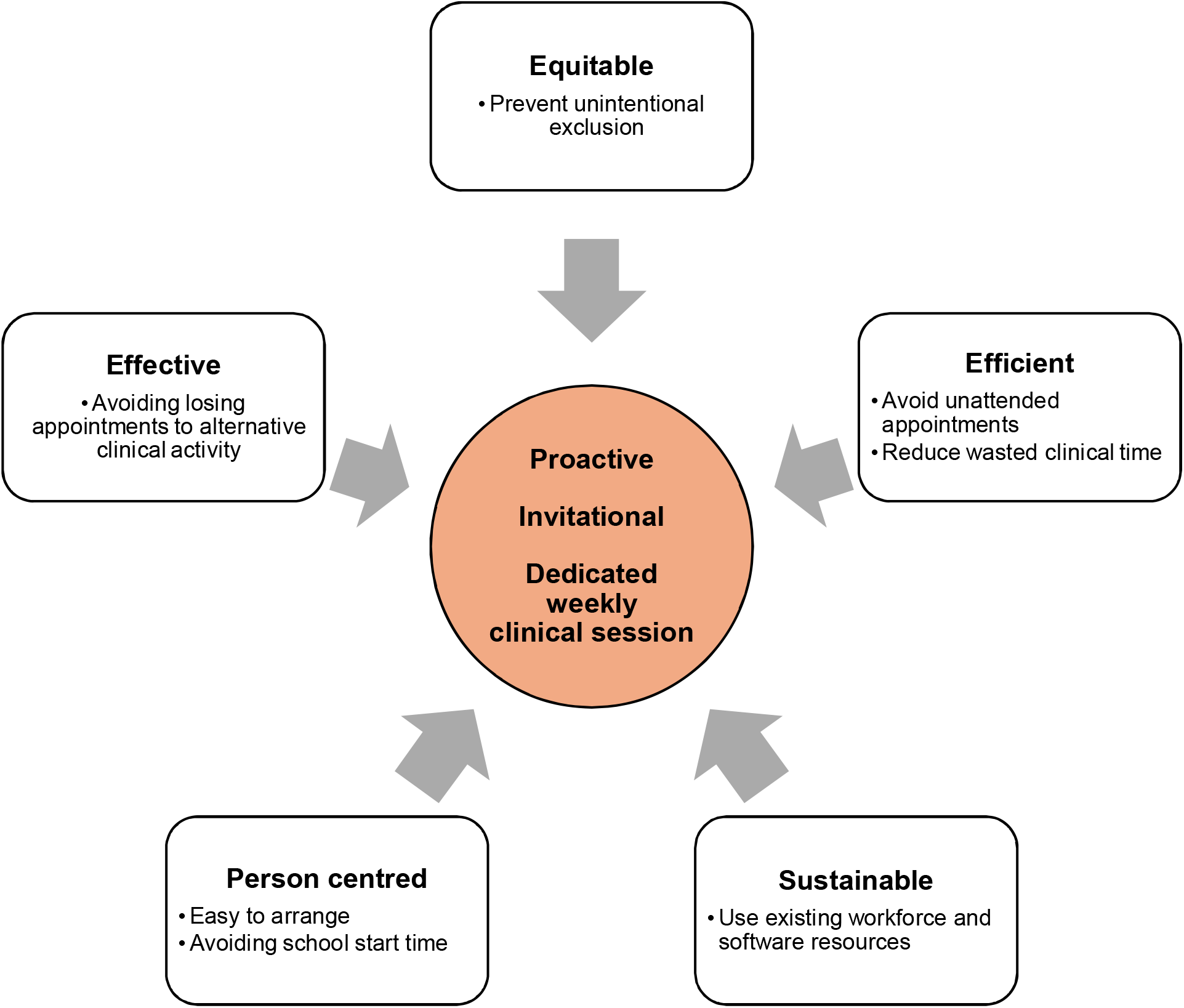

We identified three core properties for a model of access that would fulfil our priorities:

- proactive invitation of all eligible women;
- protected face-to-face appointments i.e. appointments that could not be used for alternative clinical activity;
- dedicated weekly clinics to deliver the appointments.

A practice protocol was developed whereby following receipt of a maternity discharge letter or registration of a baby, patients were contacted via SMS using Accrux(22) software and offered a pre-booked appointment date and time. An open text response to the message offered the opportunity for confirmation or rearrangement of the appointment.

A weekly 150 minutes clinical session was dedicated to maternal and baby postnatal checks divided into 15-minute appointments. This allowed the booking of multiple births without compromising dedicated time for mother or babies. This produced a capacity for up to 16 appointments per month. Appointments could only be booked by a dedicated practice administrator and released for alternative clinical activity once appointments remained unused on the day of the clinic. A single dedicated clinician was responsible for delivering the clinic with alternative cover provided by other practitioners when required. Attendance at the appointment was prioritised over timing. Patients who missed their appointments were offered up to two additional appointments. If the date and time of the appointment was unsuitable, we endeavoured to book the appointments at an alternative time suitable for the patient.

The protected appointments were introduced in April 2023. The clinic was continuously audited following its initiation to identify potential barriers to access and learning was shared to sustain improvement.

The intervention successfully improved practice performance in the provision of mother and baby checks. A total of 112/114 (98%) of eligible women were offered a postnatal check appointment. After 12 months, we saw improved uptake of maternal postnatal checks from 58% (70/121) to 89% (101/114) and appointments performed within 6-8 weeks improving from 59% (42/70) to 76% (77/101). Seven women accepted offered appointments then cancelled their appointments. Four women did not respond to the maximum of three appointment invitations. Figure 6 shows the age distribution of the post intervention population.

**Figure 6.**
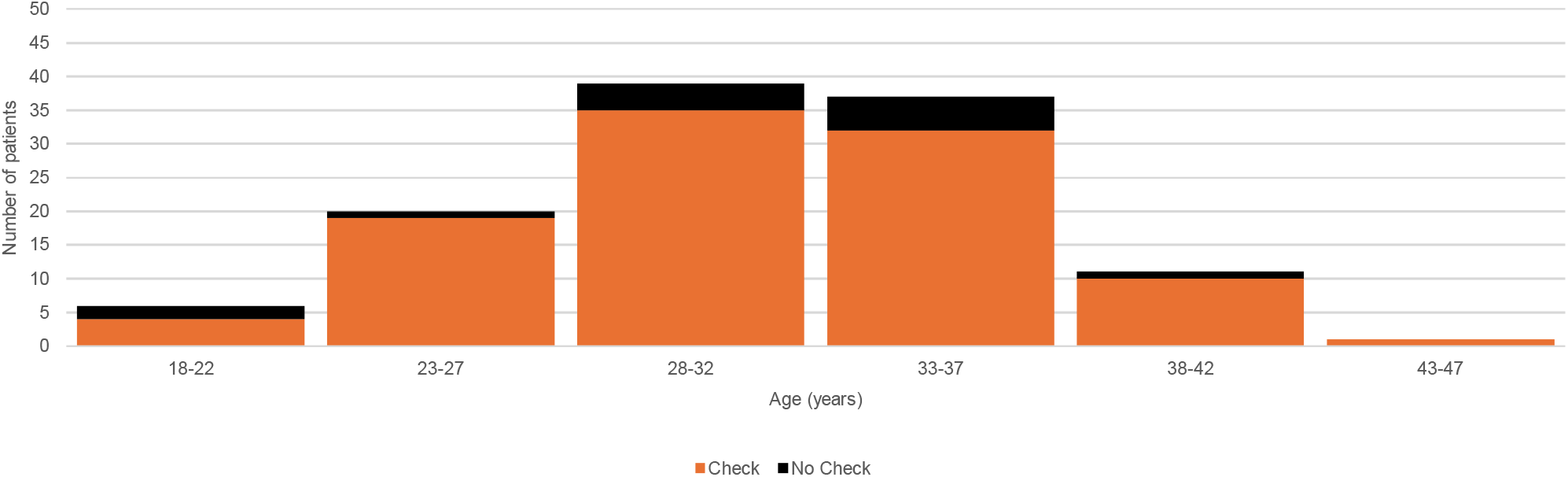
Histogram - Age distribution of post-intervention population

The uptake of newborn checks improved from 86% (113/132) to 91% (106/116) and appointments performed within 6-8 weeks improved from 46% (52/113) to 75% (80/106) [see Figure 7].

**FIGURE 7.**
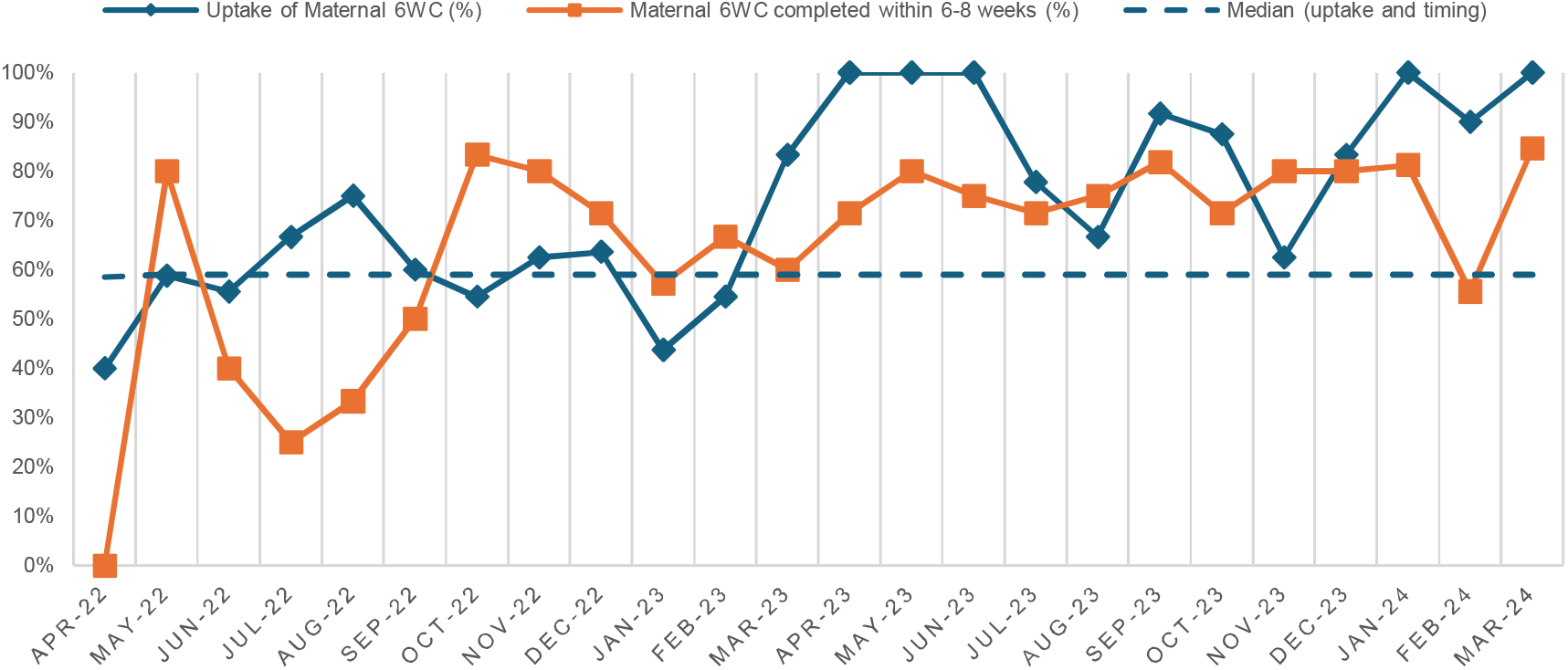
RUN CHART - MATERNAL 6WC

**FIGURE 8.**
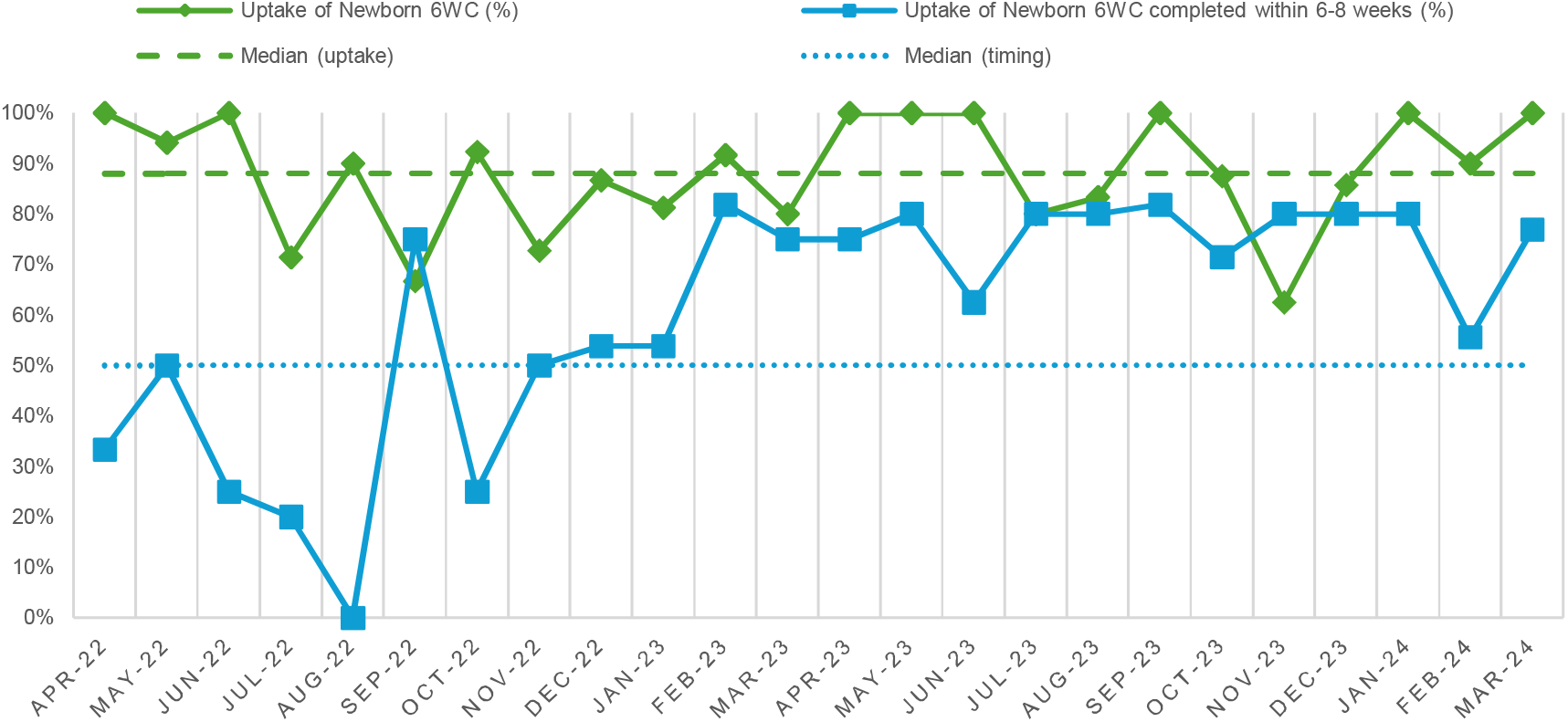
RUN CHART - NEWBORN 6WC

We identified three administrative difficulties that lead to delays in care for mothers and newborns:

- late receipt of discharge paperwork from secondary care;
- delayed registration of newborns to the practice;
- and transfer of care of babies to our practice after they were born.

## DISCUSSION

### Summary

We implemented protected postnatal appointments with proactive invitation via SMS and demonstrated sustainable improvement in service delivery over 12 months of implementation. The intervention was successful in achieving the aim of improving postnatal maternal 6WC provision without compromising newborn 6WC provision and uptake. Our aim to invite all eligible women to counter inequity resulted in 98% of eligible women being offered an appointment. The intervention required no additional workforce resources, had a low administrative burden and used digital communication tools easily available to general practices nationwide. Our intervention provides a model of access for the equitable provision of maternal postnatal care in general practice. Additionally, our intervention matched NHS England guidance introduced in December 2023, recommending that consultations must be a separate appointment and that women should be sent an invitation to the consultation(23).

### Strengths and limitations

When designing our protocol from a patient perspective, the use of SMS allowed those with reduced digital literacy to engage with their care without the use of additional software or device requirements beyond a mobile phone with SMS capabilities. As the SMS offer was the primary method for communicating appointment offers to patients, it is presumed that its use contributed to the improved uptake of appointments. It is unlikely that uptake was influenced by other factors as there were no other changes to our practice access methods. An additional strength of this protocol was that it required no new staff training as the organisation of care and use of practice resources sat within their existing skills and capabilities. A weakness of using SMS messages as a communication method is that they can produce a barrier to groups at risk of experiencing digital exclusion which can further contribute to existing heath inequalities(24). In the small number of patients this was identified for, this was mitigated by using alternative communication methods and liaising with allied health professionals.

General practitioners have professional and statutory duties to safeguard children and patterns of non-attendance may reflect additional support needs(25–27). Weekly clinics allowed recurrent lack of attendance and possible barriers to attendance to be recognised and acted upon. An additional strength of the model included it supporting the invitation of women who had experienced stillbirth or neonatal death and women whose baby remained in neonatal care by the time of the invitation.

Whilst the method of audit was time-intensive, it accurately reflected the local needs of the practice. We reduced the risk of sampling bias by capturing all women registered to the practice however data was not captured for woman who were unregistered at the time of the search. The pre-intervention audit highlighted accurate underperformance of the delivery of care. Whilst the results reflect a snapshot of a single practice, it supports the findings identified by Li et al.(2) whose study covered care delivery from 2015-2018. Code based searches relying on routinely collected data as implemented by recent studies in this area of women’s health(2,28,29) risk underestimating the provision of postnatal care by unintentionally excluding minoritised groups if data is unavailable. Patient ethnicity data for our data sets was incomplete therefore comparative analysis could not be completed.

### Comparison with existing literature

Health literacy is recognised as a contributor to disparities in postnatal care(2). The Candidacy Framework can be helpful in understanding access in general practice however individuals may struggle to identify services which fulfil the constellation of needs of a postnatal woman(11). Reduced uptake in appointments in the pre-intervention group could reflect patient disinterest in postnatal care however, comparing uptake between the two groups suggests women in our population may have been unaware of postnatal maternal checks. The improved uptake of appointments demonstrated in the post intervention population reflects how candidacy can be identified through invitations.

Macdonald et al. identified the challenges for general practitioners delivering postnatal care including the inadequacy of a standard 10-minute appointment to deliver the tasks recommended within clinical guidance. Our intervention identified administrative activity that occurred outside the delivery of direct postnatal clinical care, such as additional time taken to communicate with allied health professionals, completion of correspondence and safeguarding related administrative tasks. Current funding formulas do not remediate the patient complexity and workload of practices in deprived areas(13,30). Accounting for ‘hidden workloads’ in future service models and recognising the increased demands of working with disadvantaged populations may counter narratives in general practice burnout and retention difficulties(13,31–34).

### Implications for research and practice

The postnatal check provides an excellent opportunity for women’s health, child health and public health interventions(1,3,18,35) and its value is likely underestimated from the perspective of service commissioners. Macdonald et al. reported that 91% of UK GPs described a universal postnatal check as ‘very important’ or ‘absolutely essential’(36). Our pre-intervention audit challenges the presumption of equitable care through the universal provision of the postnatal check following the changes to the GMS contract. Paraphrasing van der Scheer et al.(37), changes in service provision contracts to address inequity are not “self-implementing”.

The 2010 Marmot Review(38) introduced the concept of ‘proportionate universalism’ as a strategy to address health inequalities. The clinical expertise, facilities, infrastructure and workforce resources required to deliver postnatal care equitably, efficiently and effectively already exists within NHS general practice resources and estates. The nature and needs of postnatal care requires dedicated and protected service delivery of high-quality individualised care by using the expert generalist skillsets of GPs. Current funding models place the obligation for provision of a comprehensive postnatal six-week check on general practice despite domains of the postnatal check sitting within maternity care, sexual and reproductive health services and public health. We advocate for the maternal postnatal check to be an essential health intervention whose delivery and funding should protected within the healthcare system even in the event of significant external factors such as a pandemic.

Over the past two decades access systems in healthcare have focussed on efficiency however recent explorations support taking a patient-centred approach over supply-focussed models of access(39). We prioritised an equity-based approach to redress disparities whilst considering the needs of the population with the capacity and abilities of our own workforce. We demonstrate an access model that can be effectively undertaken under the sceptre of improving health inequalities.

## Data Availability

All data produced in the present study are available upon reasonable request to the authors

## Appendices

## References

1. Smith HC, Saxena S, Petersen I. Postnatal checks and primary care consultations in the year following childbirth: an observational cohort study of 309□573 women in the UK, 2006–2016. BMJ Open [Internet]. 2020 Nov 23 [cited 2023 Jul 9];10(11):e036835. Available from: https://www.ncbi.nlm.nih.gov/pmc/articles/PMC7684667/

2. Li Y, Kurinczuk JJ, Gale C, Siassakos D, Carson C. Evidence of disparities in the provision of the maternal postpartum 6-week check in primary care in England, 2015–2018:an observational study using the Clinical Practice Research Datalink (CPRD). J Epidemiol Community Health [Internet]. 2022 Mar 1 [cited 2023 Jul 8];76(3):239–46. Available from: https://jech.bmj.com/content/76/3/239

3. World Health Organization. WHO recommendations on postnatal care of the mother and newborn [Internet]. Geneva: World Health Organization; 2014 [cited 2023 Jul 9]. 62 p. Available from: https://apps.who.int/iris/handle/10665/97603

4. McCauley H, Lowe K, Furtado N, Mangiaterra V, van den Broek N. Essential components of postnatal care – a systematic literature review and development of signal functions to guide monitoring and evaluation. BMC Pregnancy Childbirth [Internet]. 2022 May 28 [cited 2023 Jul 9];22(1):448. Available from: 10.1186/s12884-022-04752-6

5. British Medical Association, NHS England. Update to the GP contract agreement 2020/21–2023/24 [Internet]. 2020 [cited 2025 Apr 3]. Available from: https://www.england.nhs.uk/wp-content/uploads/2020/03/update-to-the-gp-contract-agreement-v2-updated.pdf

6. Knight M, Bunch K, Felker A, et al. Saving lives, improving mothers’ care: lessons learned to inform maternity care from the UK and Ireland Confidential Enquiries into Maternal Deaths and Morbidity 2019–21 (MBRRACE-UK) [Internet]. [cited 2025 Apr 3]. Available from: https://www.npeu.ox.ac.uk/assets/downloads/mbrrace-uk/reports/maternal-report-2023/MBRRACE-UK_Maternal_Compiled_Report_2023.pdf

7. Department of Health and Social Care. Women’s Health Strategy for England [Internet]. Department of Health and Social Care; 2022 Jul [cited 2023 Jul 1]. Available from: https://www.gov.uk/government/publications/womens-health-strategy-for-england/womens-health-strategy-for-england

8. England NHS. NHS England□» Core20PLUS5 (adults) – an approach to reducing healthcare inequalities [Internet]. [cited 2025 Apr 13]. Available from: https://www.england.nhs.uk/about/equality/equality-hub/national-healthcare-inequalities-improvement-programme/core20plus5/

9. NHS Greater Manchester, NHS Greater Manchester Primary Care Provider Collaborative. Greater Manchester Primary Care Blueprint [Internet]. 2023 Oct [cited 2025 Apr 10]. Available from: https://gmpcb.org.uk/wp-content/uploads/greater-manchester-primary-care-blueprint-october-2023.pdf

10. Wanat M, Hoste M, Gobat N, Anastasaki M, Böhmer F, Chlabicz S, et al. Transformation of primary care during the COVID-19 pandemic: experiences of healthcare professionals in eight European countries. Br J Gen Pract [Internet]. 2021 Aug 1 [cited 2025 Apr 10];71(709):e634.#x2013;42. Available from: https://bjgp.org/content/71/709/e634

11. Sinnott C, Ansari A, Price E, Fisher R, Beech J, Alderwick H, et al. Understanding access to general practice through the lens of candidacy: a critical review of the literature. Br J Gen Pract [Internet]. 2024 Oct 1 [cited 2025 Apr 5];74(747):e683.#x2013;94. Available from: https://bjgp.org/content/74/747/e683

12. Rethinking access to general practice: it’s not all about supply | The Health Foundation [Internet]. 2024 [cited 2025 Apr 10]. Available from: https://www.health.org.uk/reports-and-analysis/briefings/rethinking-access-to-general-practice-it-s-not-all-about-supply

13. Payne R, MacIver E, Clarke A. Do new models of primary care risk exacerbating existing inequity? Br J Gen Pract [Internet]. 2024 Oct 1 [cited 2025 Apr 3];74(747):436–7. Available from: https://bjgp.org/content/74/747/436

14. NCT (National Childbirth Trust). NCT (National Childbirth Trust). 2021 [cited 2023 Jul 8]. NCT finds a quarter of new mothers are not asked about their mental health. Available from: https://www.nct.org.uk/about-us/media/news/nct-finds-quarter-new-mothers-are-not-asked-about-their-mental-health

15. Six-week postnatal checks are failing many new mothers | Healthwatch [Internet]. [cited 2023 Jul 1]. Available from: https://www.healthwatch.co.uk/news/2023-03-14/six-week-postnatal-checks-are-failing-many-new-mothers

16. Maternal Postnatal Checks Cancelled during Covid-19. 2023 Jul 8 [cited 2023 Jul 9]; Available from: https://www.bmj.com/content/369/bmj.m2107/rr-2

17. Consumer Data Research Centre. English Indices of Deprivation 2010 and 2015 Data Pack [Internet]. Consumer Data Research Centre; 2016 [cited 2025 Apr 4]. Available from: https://data.cdrc.ac.uk/dataset/english-indices-of-deprivation-2010-and-2015-data-pack

18. National Institute for Health Care and Excellence. Postnatal Care NICE guideline [NG194] [Internet]. National Institute for Health Care and Excellence; 2021 [cited 2023 Jul 1]. Available from: https://www.nice.org.uk/guidance/ng194

19. Optum [Internet]. [cited 2025 Apr 4]. Transforming healthcare with tech, data and expertise. Available from: https://www.emishealth.com/

20. Arrington LA, Kramer B, Ogunwole SM, Harris TL, Dankwa L, Knight S, et al. Interrupting false narratives: applying a racial equity lens to healthcare quality data. BMJ Qual Saf [Internet]. 2024 May 1 [cited 2025 Apr 4];33(5):340–4. Available from: https://qualitysafety.bmj.com/content/33/5/340

21. Greater Manchester Health and Social Care Partnership. Greater Manchester Quality Improvement Framework [Internet]. 2023 [cited 2023 Jul 1]. Available from: https://www.england.nhs.uk/north-west/wp-content/uploads/sites/48/2019/03/Quality-Improvement-Framework.pdf

22. Accurx | The simple way to communicate about patient care [Internet]. [cited 2025 Apr 4]. Available from: https://www.accurx.com/

23. NHS England. GP six to eight week maternal postnatal consultation – what good looks like guidance [Internet]. 2023 [cited 2025 Apr 12]. Available from: https://www.england.nhs.uk/long-read/gp-six-to-eight-week-maternal-postnatal-consultation-what-good-looks-like-guidance/

24. Good Things Foundation. Mitigating Risks of Digital Exclusion in Health Systems [Internet]. 2024 [cited 2025 Apr 12]. Available from: https://www.goodthingsfoundation.org

25. RCGP. RCGP safeguarding standards for general practice [Internet]. [cited 2025 Apr 12]. Available from: https://www.rcgp.org.uk/learning-resources/safeguarding-standards

26. Greater Manchester Safeguarding Children Partnerships. Greater Manchester Safeguarding Children Procedures Manual [Internet]. 2024 [cited 2025 Apr 12]. Available from: https://greatermanchesterscp.trixonline.co.uk/chapter/using-this-manual

27. NSPCC Learning [Internet]. [cited 2025 Apr 12]. Why language matters: digging deeper than ‘did not attend’. Available from: https://learning.nspcc.org.uk/news/why-language-matters/digging-deeper-than-did-not-attend

28. Smith HC, Schartau P, Saxena S, Petersen I. The first 100 days after childbirth: cross-sectional study of maternal clinical events and health needs from primary care. Br J Gen Pract [Internet]. 2024 Sep 1 [cited 2025 May 6];74(746):e580.#x2013;6. Available from: https://bjgp.org/content/74/746/e580

29. Zhang CX, Quigley MA, Bankhead C, Kwok CH, Parekh N, Carson C. Ethnic inequities in 6–8 week baby check coverage in England 2006– 2021: a cohort study using the Clinical Practice Research Datalink. Br J Gen Pract [Internet]. 2024 Sep 1 [cited 2025 May 6];74(746):e595.#x2013;603. Available from: https://bjgp.org/content/74/746/e595

30. Blane D, Lunan C, Bogie J, Albanese A, Henderson D, Mercer S. Tackling the inverse care law in Scottish general practice: policies, interventions and the Scottish Deep End Project [Internet]. University of Glasgow, University of Edinburgh; 2024 Mar [cited 2025 Apr 12]. Available from: https://www.gla.ac.uk/media/Media_1063909_smxx.pdf

31. Woolford SJ, Watson J, Reeve J, Harris T. The real work of general practice: understanding our hidden workload. Br J Gen Pract [Internet]. 2024 May 1 [cited 2025 Apr 12];74(742):196–7. Available from: https://bjgp.org/content/74/742/196

32. Hutchinson J, Gibson J, Kontopantelis E, Checkland K, Spooner S, Parisi R, et al. Trends in full-time working in general practice: a repeated cross-sectional study. Br J Gen Pract [Internet]. 2024 Oct 1 [cited 2025 Apr 12];74(747):e652.#x2013;8. Available from: https://bjgp.org/content/74/747/e652

33. Dumast L de, Moore P, Snell KI, Marshall T. Trends in clinical workload in UK primary care 2005–2019:a retrospective cohort study. Br J Gen Pract [Internet]. 2024 Oct 1 [cited 2025 Apr 12];74(747):e659.#x2013;65. Available from: https://bjgp.org/content/74/747/e659

34. Parisi R, Lau YS, Bower P, Checkland K, Rubery J, Sutton M, et al. GP working time and supply, and patient demand in England in 2015–2022:a retrospective study. Br J Gen Pract [Internet]. 2024 Oct 1 [cited 2025 Apr 12];74(747):e666.#x2013;73. Available from: https://bjgp.org/content/74/747/e666

35. Essential Interventions, Commodities and Guidelines for Reproductive, Maternal, Newborn and Child Health [Internet]. World Health Organization (WHO); 2012 [cited 2023 Jul 9]. Available from: https://www.who.int/publications-detail-redirect/essential-interventions-commodities-and-guidelines-for-reproductive-maternal-newborn-and-child-health

36. Macdonald C, Cross-Sudworth F, Quinn L, MacArthur C, Bick D, Jones E, et al. Content and timing of the 6-8 week maternal postnatal check: a mixed methods study. BJGP Open [Internet]. 2024 Sep 30 [cited 2025 Apr 4]; Available from: https://bjgpopen.org/content/early/2024/09/25/BJGPO.2024.0229

37. van der Scheer JW, Woodward M, Ansari A, Draycott T, Winter C, Martin G, et al. How to specify healthcare process improvements collaboratively using rapid, remote consensus-building: a framework and a case study of its application. BMC Med Res Methodol. 2021 May 11;21(1):103.

38. Michael Marmot, Peter Goldblatt, Jessica Allen, et al. Fair Society, Health Lives: The Marmot Review [Internet]. 2010 [cited 2025 Apr 10]. Available from: https://www.instituteofhealthequity.org/resources-reports/fair-society-healthy-lives-the-marmot-review/fair-society-healthy-lives-full-report-pdf.pdf

39. Eccles A, Bryce C, Driessen A, Pope C, MacLellan J, Gronlund T, et al. Access systems in general practice: a systematic scoping review. Br J Gen Pract [Internet]. 2024 Oct 1 [cited 2025 Apr 3];74(747):e674.#x2013;82. Available from: https://bjgp.org/content/74/747/e674

